# Taught but not retained: disaster medicine in the education of seven health professions in Germany—a nationwide cross-sectional survey of 8,133 students and trainees

**DOI:** 10.64898/2026.10.04.26363870

**Authors:** Klaas F. Franzen, Markus Ries, Timo Ziegmann, Vincent Göhren, Benjamin Gleitz, Florian Fischer, Victoria Edwards, Amelie Klapproth, Erik Schneider, Bernhard Elsner, Robert Wunderlich, Peter Bradl, Andreas Follmann, Laura Over-Müller, Wiebke Zweig, Peer Franzen, Kristina Büscher, Daniel Drömann, Ole Franzen, Kai Mortensen, Michael Reppel, Patrick Ristau, Andrea C. Franzen, Maik von der Forst

**Affiliations:** University of Lübeck, Medical Clinic III, 23562 Lübeck, Germany; Airway Research Center North, Member of the German Center for Lung Research (DZL), 22927 Großhansdorf, Germany; Reinfeld Volunteer Fire Brigade, City of Reinfeld, 23858 Reinfeld, Germany; Heidelberg University, Medical Faculty Heidelberg, Crisis and Disaster Management Unit, Im Neuenheimer Feld 420, 69120 Heidelberg, Germany; Heidelberg University, Medical Faculty Heidelberg, Pediatric Neurology and Metabolic Medicine, Center for Pediatrics and Adolescent Medicine, Im Neuenheimer Feld 430, 69120 Heidelberg, Germany; Rhine-Neckar Public Health Service, Kurfürsten-Anlage 38–40, 69115 Heidelberg, Germany; University of Lübeck, Department of Health Sciences, Physiotherapy, 23562 Lübeck, Germany; Fraunhofer Research Institution for Individualized Medical Technology and Engineering IMTE, 23562 Lübeck, Germany; University Department of Anesthesiology and Intensive Care Medicine, University Hospital Tübingen, Eberhard Karls University Tübingen, Hoppe-Seyler-Str. 3, 72076 Tübingen, Germany; German Society for Disaster Medicine (Deutsche Gesellschaft für Katastrophenmedizin e.V.), Kirchseeon, Germany; Institute of Rescue Engineering, Emergency, Disaster Management, Würzburg-Schweinfurt Institute of Technology, Südwestpark 37-41, 90449 Nürnberg, Germany; University Hospital RWTH Aachen, Medical Faculty RWTH Aachen, Department of Anesthesiology, Pauwelsstr. 30, 52074 Aachen, Germany; Hamburg Fire Brigade, Ministry of the Interior and Sport of the Free and Hanseatic City of Hamburg, 20099 Hamburg, Germany; University of Lübeck, Medical Clinic I, 23562 Lübeck, Germany; Stormarn technical operations management, Stormarn district fire brigade association, 23843 Travenbrück, Germany; Ministry of the Interior, Municipal Affairs, Housing and Sports, 24105 Kiel, Germany; Cardiology Kiel and University of Lübeck, Department of Rhythmology, 24103 Kiel, Germany; Cardiology Landsberg and University of Lübeck, Department of Rhythmology, 86899 Landsberg, Germany; Faculty of Applied Health Sciences, Deggendorf Institute of Technology, 94469 Deggendorf, Germany; Schön Klinik Neustadt (i. Holstein), Department of Anesthesiology and Intensiv Care Medicine, 23730 Neustadt, Germany; Heidelberg University, Medical Faculty Heidelberg, Department of Anesthesiology, Im Neuenheimer Feld 420, 69120 Heidelberg, Germany

**Keywords:** Disaster medicine, health professions education, curriculum evaluation, interprofessional education, knowledge retention, willingness to respond, cross-sectional survey, Germany

## Abstract

**Background:** Disaster medicine has been a formal curricular recommendation in German health professions education since 2006, and comparable recommendations exist internationally. Evidence on implementation is confined almost entirely to human medicine, rests on faculty self-reports or small student samples, and measures delivery rather than retention. Whether curricular exposure is associated with willingness to serve has not been adequately examined in interprofessional samples.

**Objective:** We aimed to characterize disaster medicine education across 7 health professions in Germany, distinguishing delivery from retention, to identify profession-specific gaps and training priorities, and to examine how educational and experiential factors relate to willingness to serve.

**Methods:** We conducted an anonymous nationwide online cross-sectional survey of students and trainees across 7 health professions in Germany between July 9 and August 25, 2026, recruited through institutional distribution lists. The interprofessionally developed instrument comprised 48 items. We assessed delivery and retention of 12 disaster medicine topics (4-point scale), self-rated confidence in 22 clinical and nursing skills (6-point scale), desired content, formats, motivators and barriers, and formal and spontaneous willingness. For all rating scales, higher values denote the more favorable level. We derived a delivery-retention gap and a want-deficit index, examined the dimensional structure of the competence battery, tested for differential item functioning by prior civil protection engagement, and modeled willingness with a cumulative link model.

**Results:** Of 8133 respondents (completion rate 87.5%), 33.1% had never been taught disaster medicine and 25.0% only in passing; 22.1% had comprehensive teaching including practical exercises. Across the 12 topics, the proportion able to retrieve content confidently never exceeded 23.9%; the delivery-retention gap reached 47.3 percentage points. More than half (54.6%) had never taken part in a practical exercise. Competence formed more than 1 dimension: 18 acute-technical items loaded 0.54-0.86 on the first factor, 4 relational and nursing items 0.14-0.28. Prior civil protection engagement was associated with higher confidence on every acute-technical item (Cliff delta 0.26-0.55) and lower confidence on all 4 relational items (−0.22 to −0.09). Willingness was high and homogeneous across groups (78.7% formal, 84.1% spontaneous), whereas competence varied by about 2 scale points. Adjusted, active civil protection engagement (odds ratio 2.49, 95% CI 2.12-2.94) and practical exercise experience (odds ratio 1.78, 95% CI 1.54-2.05) were the strongest predictors; curricular exposure contributed weakly (odds ratio 1.06 per SD, 95% CI 0.99-1.14).

**Conclusions:** Two decades after the curricular recommendation, disaster medicine is largely absent from health professions education in Germany, and what is delivered is not retained. Willingness is nevertheless high and largely decoupled from competence, so qualification and recruitment require separate instruments. Curricula should be prioritized by the coincidence of demand and deficit rather than by coverage, built for retention, and should treat relational and nursing competence as a distinct learning objective.

## Introduction

Preparing health professionals for situations in which routine resources are insufficient is an educational problem that can become an operational one. The competencies required in a major incident, prolonged power outage, pandemic, or state of national defense differ from those required in individual patient care, and they cannot be improvised at the moment of need. Personnel is the one element of surge capacity that cannot be procured, stockpiled, or built at short notice ^1–3^. In Germany, this has moved from a theoretical to a planning-relevant concern: the framework directive on total defense adopted in 2024 foresees extensive involvement of the civilian health system, and the federal medical service explicitly names short intensive courses to establish basic competencies for auxiliary and support activities ^4,5^. International bodies have argued for two decades that these competencies belong in undergraduate health professions education. In Germany, a concept curriculum for disaster medicine in undergraduate teaching was published in 2006 and recommended for implementation by the Medical Faculty Council in 2007 ^6^. Evidence on whether this recommendation has taken effect is thinner than its age suggests and is limited in three ways. First, it is almost exclusively confined to human medicine. Two principal German studies surveyed 992 medical students nine years after the curriculum was approved and 25 of 36 medical faculties, respectively, both concluding that disaster medicine remains underrepresented and institutionally heterogeneous ^7,8^. Comparable national surveys from the Netherlands and Italy have reported similar results for medical students ^9,10^. Nursing, therapeutic professions, midwifery science, and emergency medical service training, which together educate considerably more people than medicine, are addressed by intervention literature concerned with the effectiveness of individual training formats rather than by any status quo assessment ^11^.

Second, existing evidence measures delivery, rather than retention. Faculty surveys record whether a topic appears in the curriculum and student surveys record self-assessed knowledge at a single point in time. Neither design distinguishes content that was never taught from content that was taught and has since been lost, although the educational consequences of the two are entirely different: the former is a curriculum gap, and the latter is a spacing, consolidation, or assessment issue.

Third, the literature on curricula and willingness to serve has developed separately. A systematic review of 37 studies covering 11,168 medical students found a weighted mean willingness to volunteer of 68.4% alongside poor disaster knowledge and minimal curricular coverage, and documented a significant discrepancy between planned and actual volunteering ^12^. The coexistence of both deficits is well established. Whether they are causally or associatively linked, that is, whether teaching disaster medicine makes students more willing to serve, has not been tested in an adequately powered sample with an adjusted model. The assumption that it does underlies most calls for curricular expansion, including our own initial expectations.

These three limitations are not independent of each other. They persist together because the necessary study must be interprofessional, large enough to model group differences and adjusted associations simultaneously, and instrumented to separate delivery from retention. The present study was designed to address these issues. We conducted a nationwide survey of students and trainees across seven occupational groups and asked four questions to 1) characterize the current state of disaster medicine education, 2) identify profession-specific gaps and 3) training priorities, and 4) examine how educational and experiential factors relate to willingness to serve

1. What is the actual status of disaster medicine content in health professions education in Germany, distinguishing between delivery and retention?
2. How do these statuses and self-assessed competencies differ between occupational groups?
3. What content do learners want, and where does that demand coincide with the greatest deficit?
4. Which characteristics predict individual willingness to be recruited and trained once sociodemographic and occupational characteristics are held constant?

This analysis forms part of a nationwide project in Germany that surveyed both students and trainees and the organizations that would deploy them using item-for-item mirrored instruments. The alignment between the two perspectives is reported in a companion study ^13^. This study focuses solely on the education sector.

## Methods

### Design

Anonymous, non-interventional, nationwide, online, cross-sectional survey in Germany. Reporting follows STROBE for cross-sectional studies and CHERRIES for internet-based surveys in its corrected version ^14–16^, and dispositions follow the AAPOR Standard Definitions ^17^. Both checklists can be obtained from the corresponding author upon request. The analysis and publication plans are available online ^13^.

### Populations and Eligibility

Eligible participants were those enrolled in a course of study or vocational training in a health, nursing, therapeutic, or emergency medical service occupation in Germany, aged 18 years or older, with sufficient German language proficiency, who provided active consent, and processed at least the first substantive question block. Eligible programs covered human and dental medicine, psychology and psychotherapy, nursing in both degree and vocational routes, physiotherapy, occupational therapy, speech and language therapy, midwifery science, physician assistance, paramedic and emergency medical technician training, and medical-technical training. Individuals under 18 years were excluded by design because the instrument addresses willingness to expose oneself to potentially hazardous and distressing deployment situations, and involving legal guardians would have required identification of participants and was therefore incompatible with the anonymous design.

For the analysis, programs were aggregated into seven occupational groups fixed before analysis: human medicine, nursing, emergency medical services, therapeutic professions, midwifery and physician assistance, psychosocial professions, and others (Table 2). One questionnaire coding issue was identified: two response options carried identical texts under different codes and were merged in every code-based analysis.

### Recruitment and Field

Recruitment proceeded exclusively through institutional distribution lists and publicly accessible channels, including student representative bodies, university and school distribution lists, professional associations, and learned societies. No named individuals were directly approached by the study team. Onward forwarding by recipients was possible and not controlled. Up to two reminder waves were sent through the same channel. The field was run from July 9 to August 25, 2026. Because the survey was open and no sampling frame existed, no response rate in the strict sense was calculable; we report the completion rate as defined in the corrected version of CHERRIES ^14,15^.

### Instrument and Derived Measures

The instrument was developed specifically for this project and consented across four development stages by an interprofessional expert panel drawn from emergency and disaster medicine, nursing, emergency medical services, higher education, and civil protection, followed by a cognitive pretest for comprehensibility, processing time, and filtering logic. Neither the panel feedback nor the pretest responses were included in the analysis. The instrument was implemented on a LimeSurvey installation hosted in Germany, comprising nine pages and 48 questions, with a median processing time of 13.5 min. A completeness check prompted respondents about unanswered items before submission, but only consent and the confirmation of legal adulthood were mandatory; therefore, respondents could proceed regardless, which is the origin of the item non-response handled separately in Section 2.6. The instrument was not externally validated; content validity rested on the consenting process and the pretest, and internal consistency and structure were determined here for the first time and reported as exploratory.

Four derived measures require further definition.

- **Curricular exposure (what was taught and how well it was retained):** For each of the 12 disaster medicine topics (Table 1), respondents indicated whether the topic had not been taught, had been taught but was barely retained, had been taught and was partly retained, or had been taught and could be retrieved confidently. The curricular exposure score was the mean across the 12 items, ranging from 1 to 4. Higher values therefore denote greater curricular exposure and retention. The same polarity applies to the competence scale described below, so that in all tables a higher value denotes the more favorable level.
- **Delivery–retention gap (what was taught but is no longer confidently retrievable):** For each topic, the difference in percentage points between the proportion reporting that the topic was taught in any form and the proportion that could confidently retrieve it. This separates the curriculum gap from a retention problem, which the conventional delivery indicator conflates.
- **Want–deficit index (what learners want but cannot confidently retrieve):** For each topic appearing both in the list of desired additional content and in the curricular battery, the difference in percentage points between the proportion of participants desiring the content and the proportion able to confidently retrieve the corresponding curricular topic. High values identify topics that are both in demand and underserved. The assignment of the 11 topic pairs was fixed in the Codebook.
- **Self-assessed competence (how confident participants feel in performing relevant skills):** Confidence in 22 clinical and nursing skills was assessed on a six-point scale, recoded throughout, such that higher values denoted greater confidence.

**Table 1.**
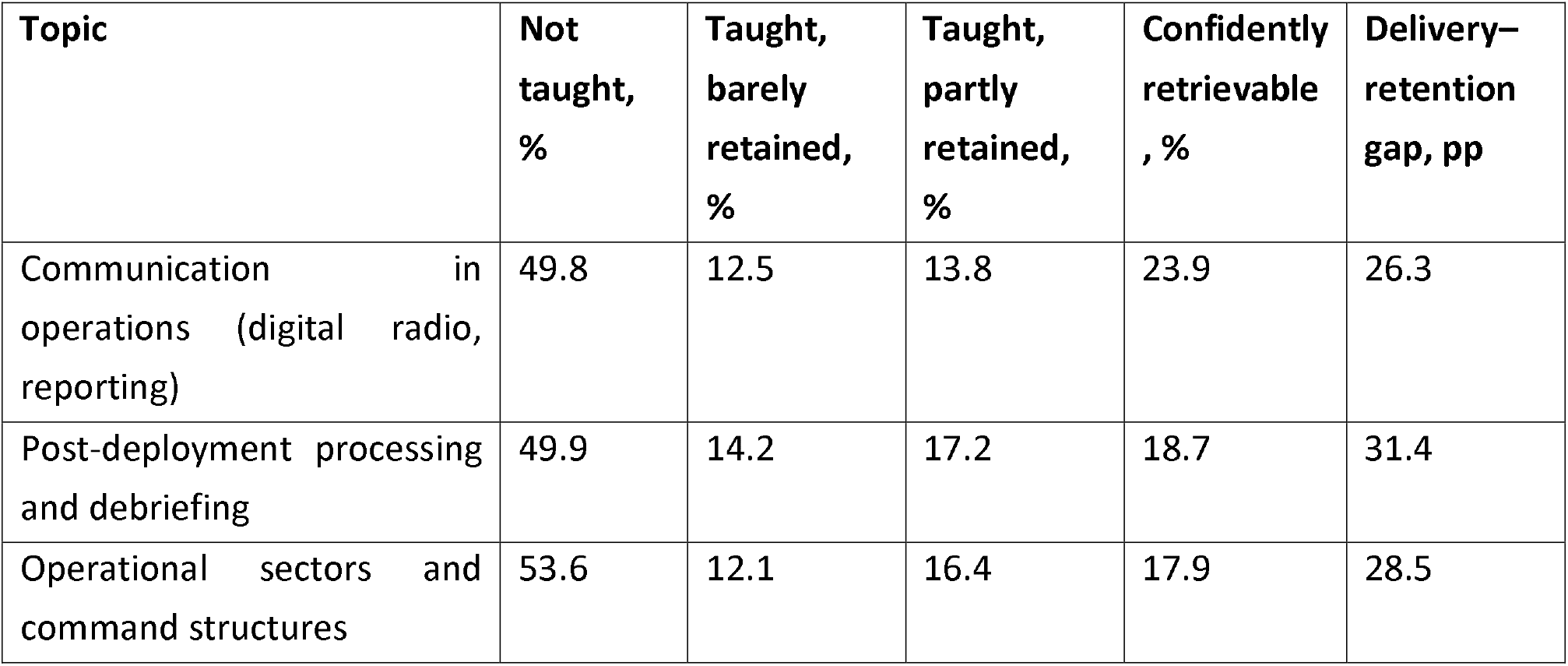
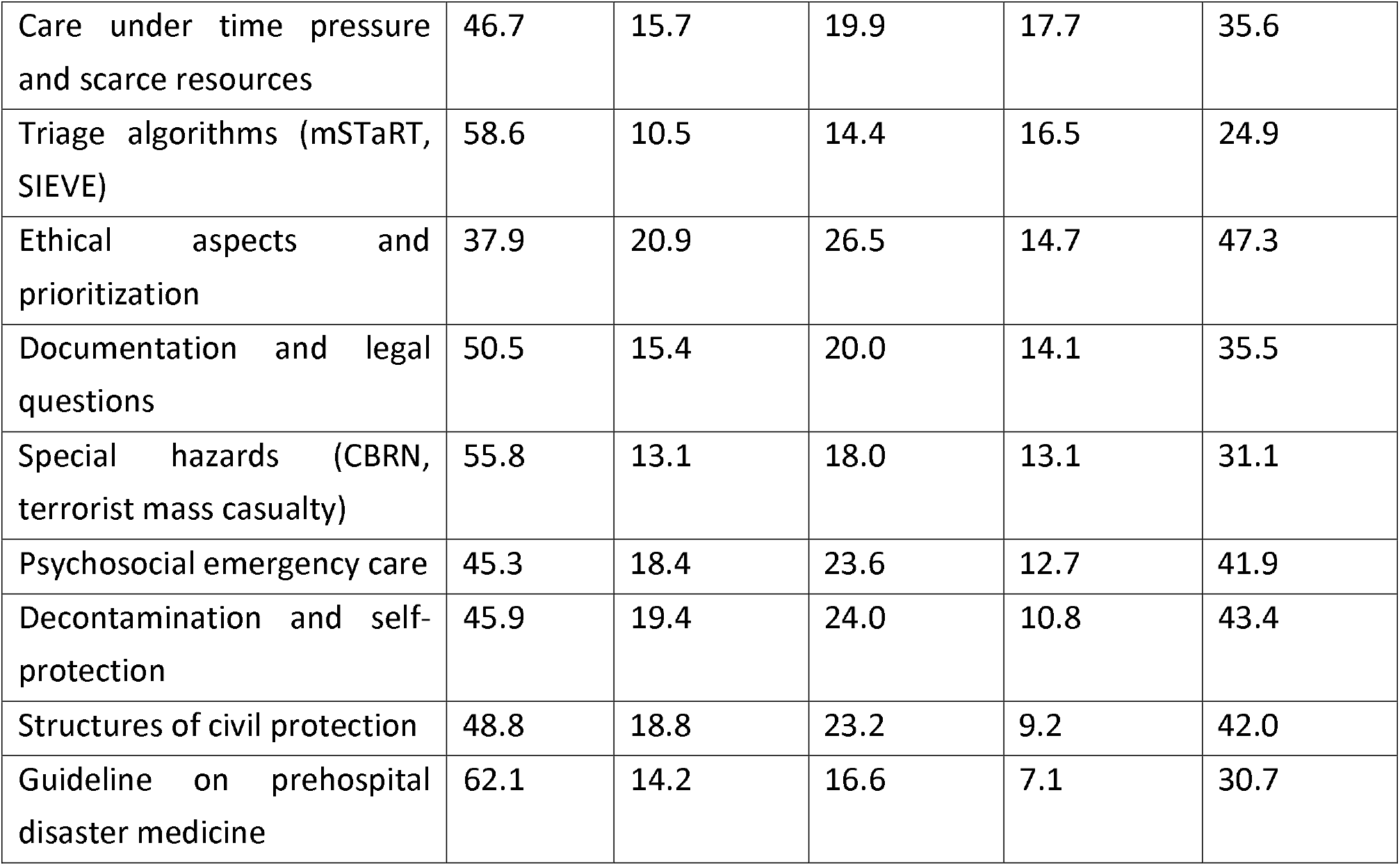
Delivery and retention of 12 disaster medicine topics (n = 7,714), ordered by the proportion of participants able to retrieve the content confidently. The delivery–retention gap is the difference between the proportion of students who report that a topic was taught in any form and the proportion of students who can confidently retrieve that information. Percentages within a row may not sum to 100 because of rounding errors.

### Endpoints

The primary endpoints were the delivery and retention of the 12 disaster medicine topics and predictors of formal willingness.

Willingness referred to stated willingness to participate in disaster response and was assessed separately as formal willingness to be recruited and trained and as willingness to provide spontaneous support if asked.

Secondary endpoints comprised self-assessed competence and its dimensional structure, the want–deficit index, desired formats and realistic time budget, framework conditions, motivators and barriers, spontaneous willingness, conceivable contributions to public health service, and the single measure considered most important by respondents.

### Statistical Analysis

The two-sided significance level was set at .05 for all analyses. All estimators are reported with 95% confidence intervals; P values supplement but do not replace the effect estimation. The full analysis set comprised all cases with consent and at least one substantive item answered with item-wise denominators. The complete set was used for the sensitivity analysis. Three sources of missing data were handled separately: filter-driven non-display, item non-response, and breakoffss. Because the denominators are formed item-wise, they vary between analyses and are reported wherever they differ from the analysis set.

Proportions were reported using Wilson score intervals (confidence intervals for binomial proportions) ^18^. Differences between occupational groups were tested using the Kruskal–Wallis test (non-parametric test for differences across multiple groups) and quantified using epsilon squared (effect-size measure). Ordinal comparisons between subgroups used Cliff delta (non-parametric effect-size measure of stochastic dominance) with bias-corrected and accelerated bootstrap intervals as a distribution-free measure of stochastic dominance ^19^. The dimensional structure of the competence battery was examined using a parallel analysis of polychoric correlations (correlations for ordinal variables) with a principal axis solution (factor-extraction method). Internal consistency was reported as Cronbach’s alpha (internal-consistency coefficient) and as a one-factor, McDonald’s omega (model-based reliability coefficient). Differential item functioning by prior civil protection engagement was assessed item-wise using Cliff delta and Benjamini–Hochberg correction (false-discovery-rate adjustment) ^20^.

Willingness was modeled using a cumulative link model with a logit link and an a priori fixed predictor set without stepwise selection, comprising age, sex, occupational group, phase of training, prior completed vocational training, civil protection engagement, radio communication training, practical exercise experience, curricular exposure score, and employment in addition to studies. The proportional odds assumption was also evaluated. Decision rules were supplemented by a region of practical equivalence of |log odds ratio| < 0.1 fixed in advance, so that a precisely estimated but negligible association was reported as such, rather than as a positive finding. Analyses labeled exploratory were controlled at a false discovery rate of 5% and were hypothesis generating. The analyses were performed in a containerized environment using fixed random seeds.

### Ethical Considerations

The study was reviewed by the Ethics Committee of the University of Lübeck (reference AZ 2026-127, March 24, 2026) in accordance with the Declaration of Helsinki; the vote preceded the commencement of data collection. Participation was voluntary and preceded by active informed consent as the first question. No names, addresses, email addresses, matriculation numbers, or IP addresses were recorded; regional information was collected only at the level of the first three postal code digits. Because no personal reference could be established, withdrawal after submission was technically impossible, and participants were informed of this in advance.

## Results

### Participants

Of the 9,375 records accessed, 9,033 provided consent, 8,133 were included in the full analysis set, and 7,119 submitted the questionnaire, resulting in a completion rate of 87.5%. The median age was 24 years (IQR 22–27), and 59.8% were women. Human medicine accounted for 33.0% of the sample, nursing for 25.7%, and emergency medical services, midwifery, physician assistance, psychosocial professions, and therapeutic professions each between 4.3% and 6.5%. Just under half (47.6%) had completed a vocational qualification in health care before entering their current program, 40.3% were actively engaged in a civil protection or response organization, and 45.4% had participated in at least one practical major incident exercise. All ten postal code lead zones were represented, with an overrepresentation of northern Germany, which is mentioned in the limitations. The full sample characteristics are given in Multimedia Appendix 1, and the field dispositions are presented in Multimedia Appendix 2.

### Disaster Medicine Is Largely Absent From the Curriculum

One-third of respondents (33.1%, 95% CI 32.1–34.2) reported that disaster medicine had not been taught at all in their program to date, and a further quarter (25.0%) reported that it had been mentioned only in passing. Comprehensive teaching, including practical exercises, was reported by 22.1% of respondents, and purely theoretical delivery by 18.8%; 1.0% did not know (n = 7,714). Nearly two decades after the German concept curriculum was recommended for implementation, a clear majority of the future health workforce has received either nothing or a passing mention.

### What Is Delivered Is Largely Not Retained

Across all 12 topics (Table 1, Figure 1), the proportion of those who could confidently retrieve the content never exceeded 23.9%, and for the clinical practice guideline on prehospital disaster medicine, it was 7.1%. The proportion of respondents who reported no deliveries ranged from 37.9% to 62.1%. For triage algorithms, arguably the single most operationally consequential topic in the list, 58.6% reported that it had never been taught at their institution. The delivery–retention gap is a more instructive quantity. It is largest for ethical aspects and prioritization (47.3 percentage points (pp)), decontamination and self-protection (43.4 pp), and the structures of civil protection (42.0 pp), that is, precisely for those topics that would be immediately action-guiding in a deployment. For these three topics, most respondents had encountered the material but could not retrieve it. The mean curricular exposure score across the twelve items was 2.00 (SD 0.71) on a scale from 1 to 4.

**Figure 1.**
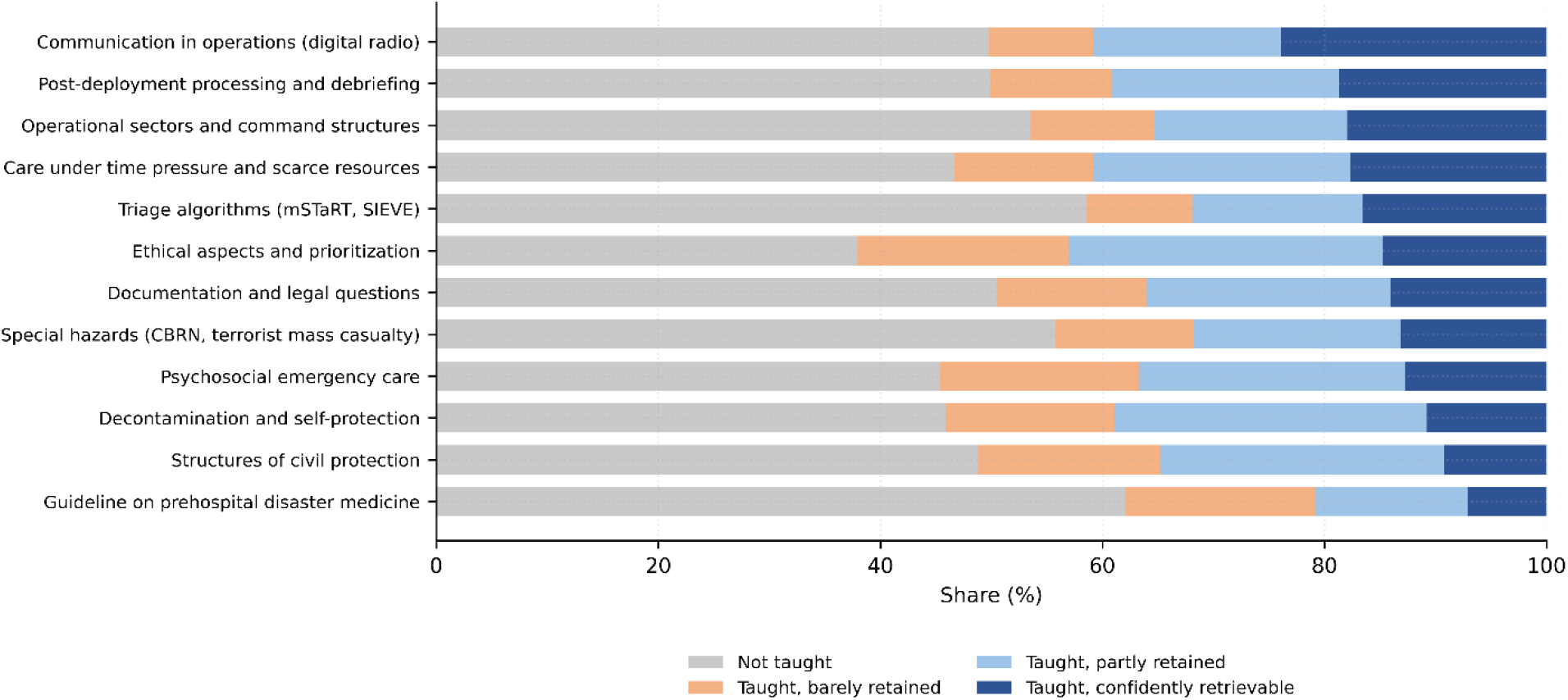
Delivery and retention across the twelve disaster medicine topics, shown as stacked proportions from “not taught” to “taught and confidently retrievable.”

### Practical Exercise Experience Is the Exception, Not the Rule

More than half of the respondents (54.6%, 95% CI 53.5–55.7) had never participated in any practical exercise or simulation related to major incidents. Where exercise experience existed, it was more often from outside the educational institution than from within it. Course participation showed the same pattern of breadth without depth: a basic first-aid course had been completed by 62.0%, but a mass casualty or triage training course by only 15.8% and a structured trauma course by 15.8% (Multimedia Appendix 3). Thus, educational institutions are not the principal providers of major incident preparation for this population.

### Occupational Groups Differ More in Competence Than in Willingness

The 7 occupational groups differed markedly in curricular exposure (Table 2), from 1.34 in the therapeutic professions to 2.88 in nursing (H=2593.0, df=6, P<.001, epsilon squared=0.346), and in self-assessed competence, from 3.36 in the psychosocial professions to 5.17 in emergency medical services (H=1766.1, df=6, P<.001, epsilon squared=0.231). Willingness differed far less: formal willingness ranged from 60.8% to 87.1% (H=227.9, df=6, P<.001, epsilon squared=0.031). Expressed on a common scale, occupational group accounted for 34.6% of the rank variance in curricular exposure and 23.1% in competence, but only 3.1% in willingness.

**Table 2.**
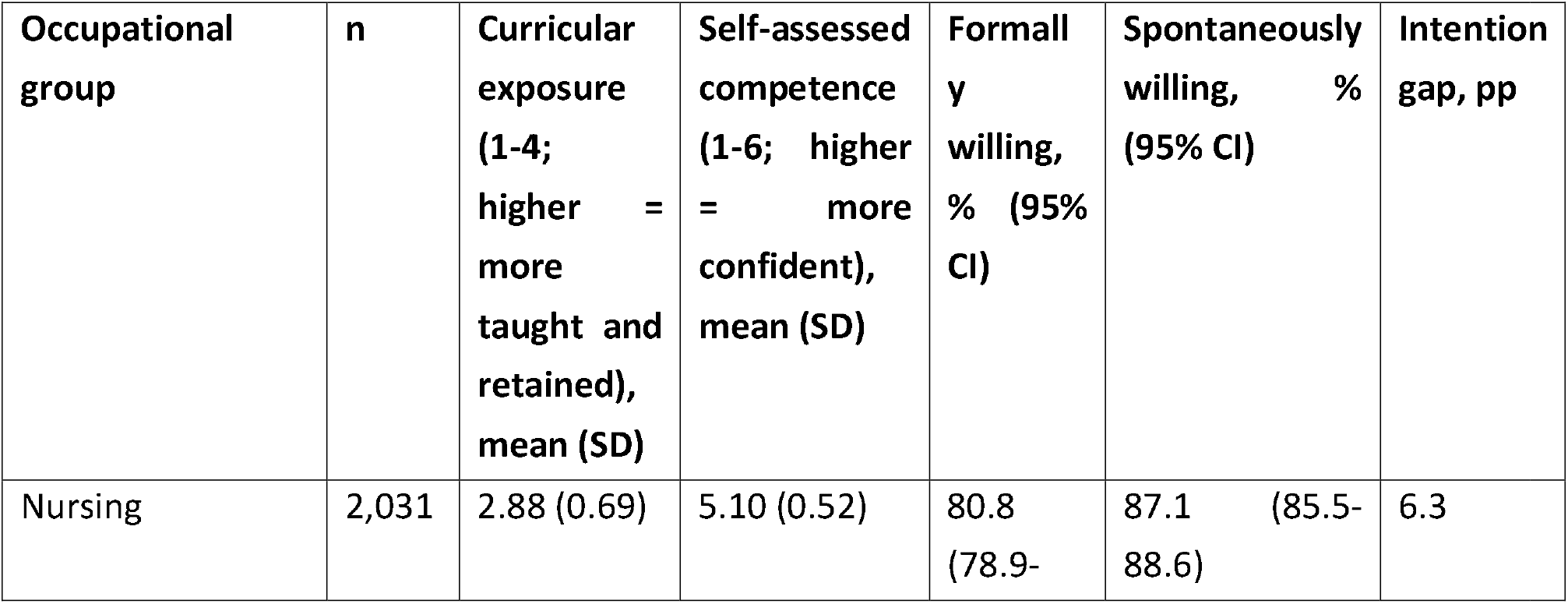
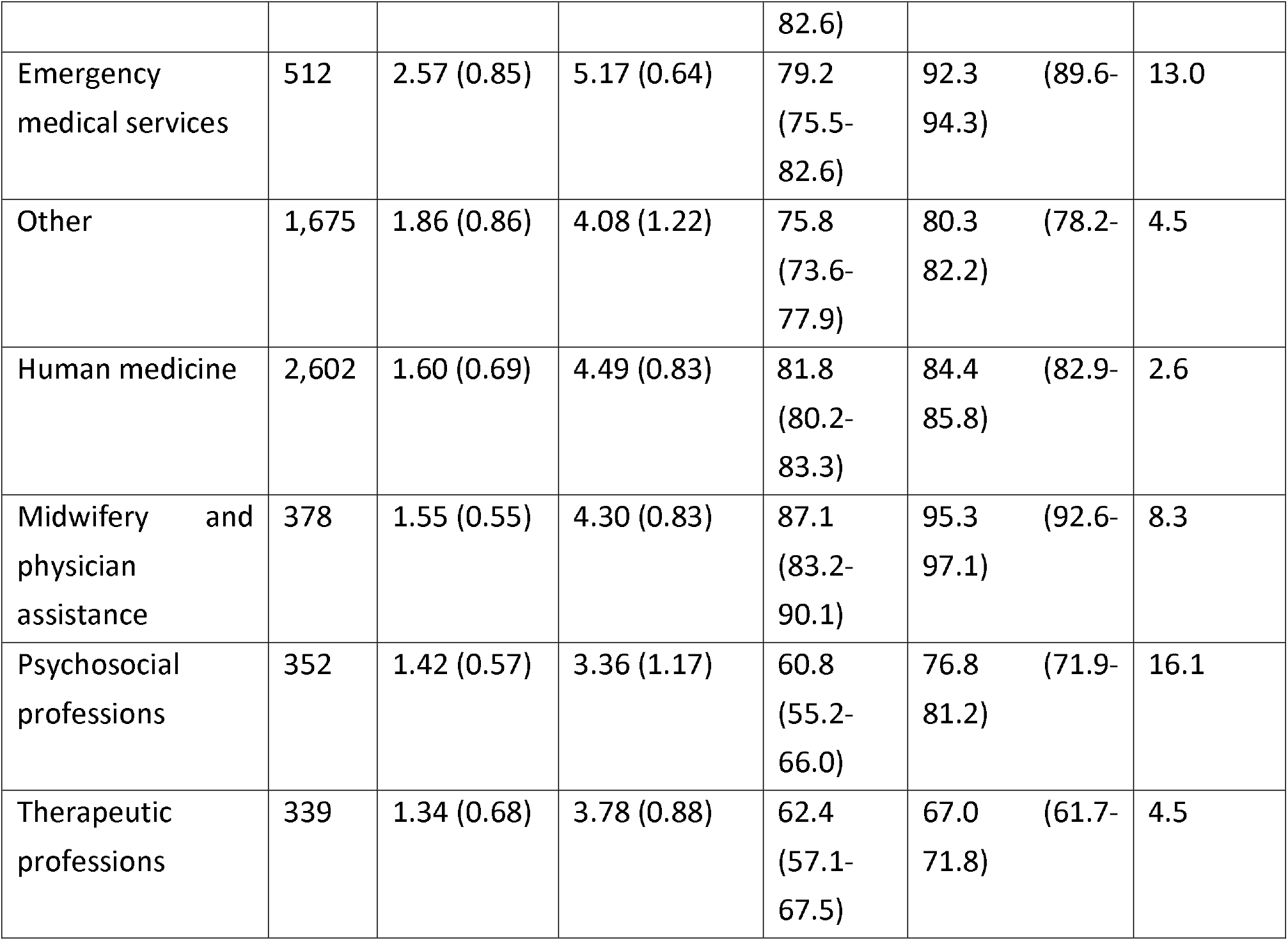
Curricular exposure, self-assessed competence, and willingness by occupational group, ordered by curricular exposure. The intention gap is the difference between spontaneous and formal willingness, measured in percentage points. For both rating scales, a higher value denotes the more favorable level: on the curricular exposure scale (1-4), 4 corresponds to “taught and confidently retrievable”; on the competence scale (1-6), 6 corresponds to “very confident.” Willingness is reported as the proportion of respondents in the top 2 response categories. Values in parentheses are SDs for the 2 rating scales and 95% Wilson score CIs for the 2 proportions. Kruskal-Wallis tests across the 7 occupational groups: curricular exposure H=2593.0, df=6, P<.001, epsilon squared=0.346; self-assessed competence H=1766.1, df=6, P<.001, epsilon squared=0.231; formal willingness H=227.9, df=6, P<.001, epsilon squared=0.031.

The two dimensions are also ordered differently in the study. Nursing and emergency medical service trainees led in terms of competence but not willingness, while midwifery and physician assistant students led in terms of willingness but sat in the lower half in terms of competence. Therefore, competence and willingness are not two expressions of the same underlying disposition, and an educational intervention that raises one should not be assumed to raise the other.

### Self-Assessed Competence Has More Than One Dimension

The competence battery showed high internal consistency (Cronbach’s alpha 0.946; one-factor McDonald’s omega 0.951), but parallel analysis retained three factors, the first accounting for 49.7% of the variance (eigenvalues 10.93, 2.99, and 1.18). The inspection of the loadings shows why. Eighteen acute-technical items loaded between 0.54 and 0.86 on the first factor, whereas four relational and nursing items loaded between 0.14 and 0.28: empathetic communication, personal care and mobilization, assistance with eating and drinking, and providing orientation in the daily routine. Therefore, a single competence construct is not tenable for this battery (see Multimedia Appendix 5).

Differential item functioning analysis makes the substantive meaning of this structure visible (Table 3). Respondents who actively engaged in civil protection reported higher confidence than the remainder on every one of the 18 acute-technical items, with Cliff delta between 0.26 and 0.55, and lower confidence on all four relational items, with Cliff delta between −0.22 and −0.09 (all Benjamini–Hochberg corrected P < .001). The reversal is complete; there is no item for which the direction is ambiguous.

**Table 3.** Differential item functioning by prior civil protection engagement: the four largest positive and negative effects. Positive values indicate higher confidence among those who are actively engaged. A complete table of all 22 skills is provided in Multimedia Appendix 6.

| Skill | Dimension | Cliff delta<br>(engaged vs. others) | 95% CI |
| --- | --- | --- | --- |
| Airway management | acute-technical | 0.55 | 0.53 to 0.58 |
| Haemorrhage control: tourniquet | acute-technical | 0.55 | 0.53 to 0.57 |
| Pre-triage in a mass casualty incident | acute-technical | 0.51 | 0.49 to 0.53 |
| Immobilization and splinting | acute-technical | 0.50 | 0.48 to 0.52 |
| Empathetic communication | relational–nursing | $-0.09$ | $-0.12$ to $-0.07$ |
| Personal care, positioning, and mobilization | relational–nursing | –0.14 | –0.16 to –0.11 |
| Assistance with eating and drinking | relational–nursing | –0.16 | –0.19 to –0.14 |
| Providing orientation in the daily routine | relational–nursing | –0.22 | –0.24 to –0.19 |

We interpret this as a substantive, rather than a psychometric, result. Exposure to civil protection structures builds acute technical confidence but does not build, and may displace, relational and nursing confidence. This pattern could not have been detected in a monoprofessional study of medical students because the relational items were absent from the instrument.

### Where Demand and Deficit Coincide

The demand for additional content was high and specific to the target audience (Table 4, Figure 2). Trauma care under resource scarcity was desired by 73.3% and could be confidently retrieved from the curriculum by 17.7%with a want-deficit index of 55.7 pp. Triage followed at 46.9 pp, and the structures of civil protection at 42.2 pp. On the other hand, digital operational tools show an index of 9.5 pp, not because they are well taught, but because they are comparatively less desired.

**Table 4.** Want–deficit index across the eleven topics present in the desired-content list and in the curricular battery. Desired content, n = 7,242; curricular battery, n = 7,714. pp=percentage points

| **Desired content** | **Desired, %** | **Corresponding curricular topic confidently retrievable, %** | **Want–deficit index, pp** |
| --- | --- | --- | --- |
| Trauma care under resource scarcity | 73.3 | 17.7 | 55.7 |
| Triage | 63.4 | 16.5 | 46.9 |
| Structures of civil protection | 51.4 | 9.2 | 42.2 |
| Crisis communication and small-team leadership | 58.5 | 17.9 | 40.5 |
| CBRN situations | 53.1 | 13.1 | 40.0 |
| Legal questions in deployment | 53.2 | 14.1 | 39.1 |
| Self-protection and self-care | 48.6 | 10.8 | 37.9 |
| Psychosocial emergency care | 45.8 | 12.7 | 33.0 |
| Dealing with affected persons and the dying | 40.1 | 14.7 | 25.4 |
| Post-deployment processing and ethics | 42.6 | 18.7 | 23.9 |
| Digital operational tools and documentation | 33.4 | 23.9 | 9.5 |

**Figure 2.**
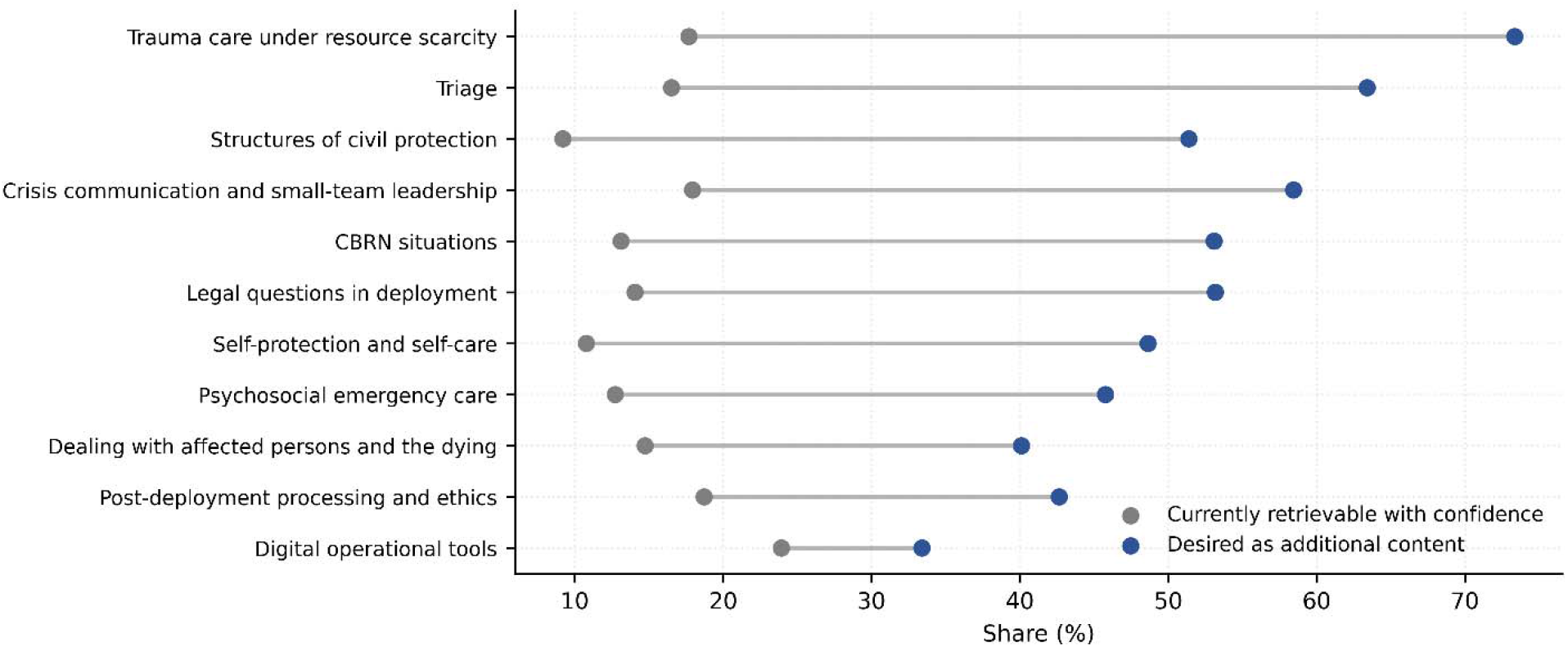
Want–deficit comparison: the proportion of participants desiring each content plotted against the proportion able to confidently retrieve the corresponding curricular topic.

The index was deliberately constructed to rank topics by the joint presence of demand and deficit rather than by either alone, and it produces a different ordering from the delivery indicator that curriculum reviews conventionally use. For instance, ethical aspects have the largest delivery–retention gap of all 12 topics but rank ninth on the want–deficit index because the demand for them is comparatively modest. Whether a curriculum should follow demand at this point is a judgment that the index makes visible rather than settling.

### Formats, Time Budget, and Conditions

The preferred formats were practical simulation days and exercises with response organizations (68.8%), elective modules (56.3%), compact in-person courses, such as weekend blocks (56.1%), and hybrid formats (54.8%). Compulsory integration into the regular curriculum was supported by 52.2% of respondents, and live online seminars by 24.7% (n = 7,242). The realistic maximum time budget was one weekend per half-year for 38.2%, up to 20 hours per semester for 22.0%, two to three days per year for 21.6%, and more than 20 hours per semester if creditable for 15.7% of the respondents. Only 1.4% of the respondents stated that no additional time was possible.

Among the framework conditions, supervision by experienced instructors ranked first (95.6%), followed by the offer being free-of-charge (93.8%) and substantial practical components (93.6%). Insurance coverage was 83.4% and a clear legal position was 81.6%. Academic credit ranked last among the eight conditions (62.5%), below a recognized certificate (72.8%). This ordering is worth noting because credit and certification are the levers curriculum designers most readily reach.

### Motivation Is Intrinsic, Obstruction Is Structural

The strongest motivators were the acquisition of practical competence (86.0% rated it as a strong motivator) and contributing to society (81.3%), followed by insurance and legal protection (73.2%) and financial compensation (66.1%). The strongest barriers were lack of insurance coverage (63.0%) and time burden of studies or training (60.4%), followed by family obligations (48.0%) and unclear legal position (47.8%). Concern about being professionally out of one’s depth ranked eighth (36.3%), and concern about psychological burden ranked last (22.7%) (Multimedia Appendix 9).

Asymmetry is informative for designing interventions. This population is attracted by competence and purpose, but obstructed by insurance, time, and legal ambiguity. None of the leading barriers are attitudinal, and none are addressed by persuasion.

### Exposure Predicts Willingness; Curricular Exposure Barely Does

Willingness was high in absolute terms: 80.9% considered systematic recruitment and training sensible, 78.7% were willing to be formally recruited and trained, and 84.1% would support spontaneously if asked (n = 7,472). Spontaneous willingness exceeded formal willingness in every occupational group by between 2.6 and 16.1 percentage points, and the cross-tabulation shows that the asymmetry is systematic rather than driven by a subgroup (Multimedia Appendix 10). Therefore, what limits formal commitment is bindingness, not helpfulness.

After adjustment, the strongest predictor was active engagement in a civil protection or response organization, followed by practical exercise experience, previous engagement, and radio communication training (Table 5, Figure 3). Curricular exposure contributed independently but weakly, with a confidence interval within the region of practical equivalence specified in advance. Prior vocational qualifications and employment alongside studies did not contribute to the results.

**Table 5.** Cumulative link model for formal willingness (n = 7,235). Odds ratios above 1 indicate a higher willingness. The full model, including the age spline, is presented in Multimedia Appendix 7.

| Predictor | OR | 95% CI | Interpretation<br>diese Spalte erscheint mir etwas uneinheitlich (Bedeutung vs. Referenzbezug) und ist daher noch nicht so gut verständlich, vielleicht sollten wir in der Table legend erklären, was wir hier aussagen möchten |
| --- | --- | --- | --- |
| Active civil protection engagement | 2.49 | 2.12 to 2.94 | strongest predictor |
| Practical exercise experience | 1.78 | 1.54 to 2.05 | — |
| Former civil protection engagement | 1.45 | 1.24 to 1.70 | — |
| Radio communication training | 1.32 | 1.16 to 1.51 | — |
| Late phase of training | 1.17 | 1.05 to 1.31 | — |
| Prior vocational qualification in health care | 1.11 | 0.98 to 1.26 | not distinguishable from no effect |
| Employment alongside studies | 1.10 | 0.98 to 1.23 | not distinguishable from no effect |
| Curricular exposure, per SD | 1.06 | 0.99 to 1.14 | within the region of practical equivalence |
| Female sex | 0.85 | 0.76 to 0.95 | — |
| Occupational group: midwifery and physician assistance | 1.55 | 1.20 to 2.00 | reference: human medicine |
| Occupational group: psychosocial professions | 0.73 | 0.58 to 0.92 | reference: human medicine |
| Occupational group: therapeutic professions | 0.61 | 0.49 to 0.77 | reference: human medicine |
| Occupational group: nursing | 0.52 | 0.45 to 0.60 | reference: human medicine |
| Occupational group: emergency medical services | 0.41 | 0.33 to 0.52 | reference: human medicine |

**Figure 3.**
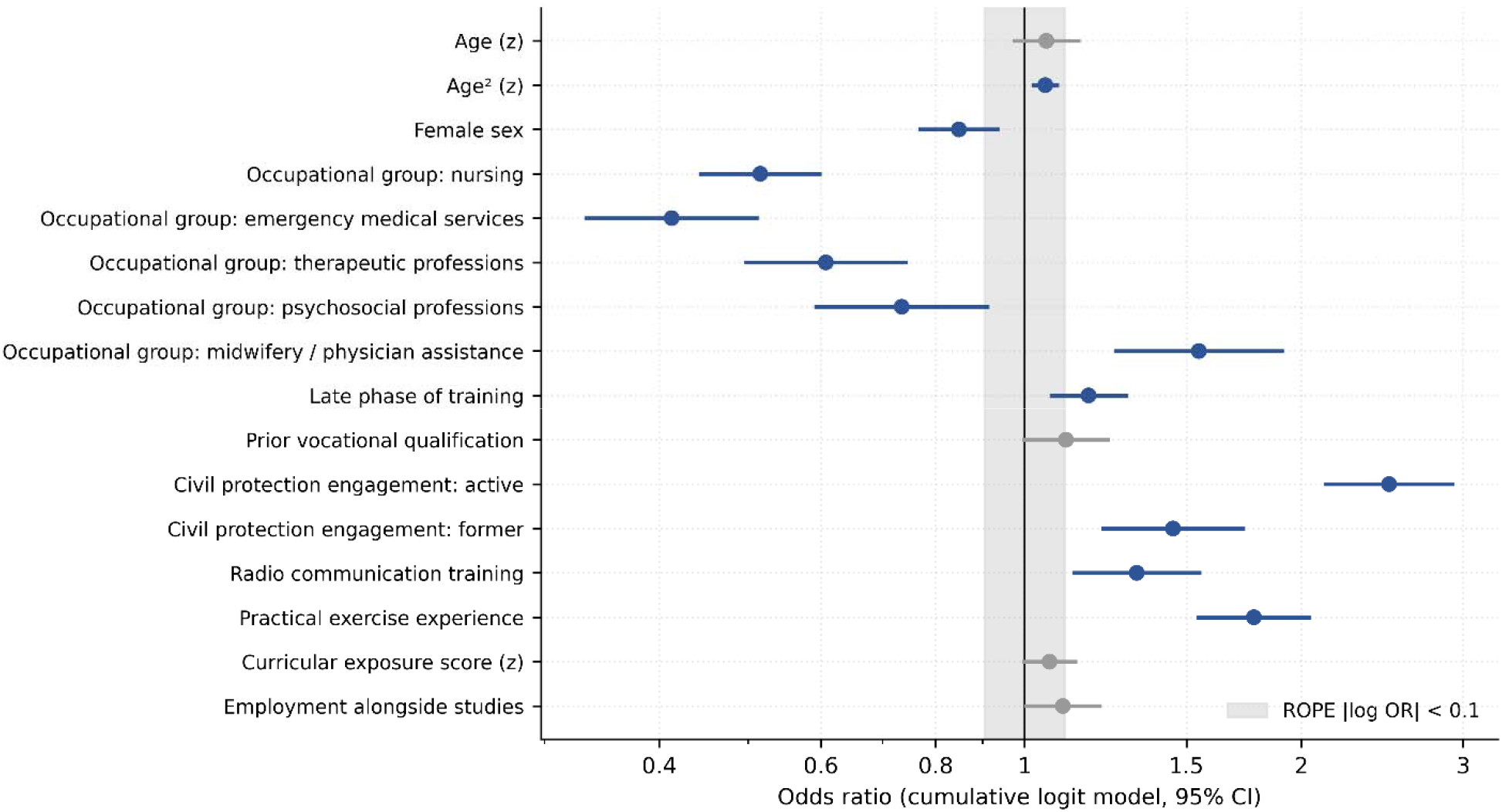
Adjusted odds ratios with 95% confidence intervals for formal willingness. The shaded band marks the pre-specified region of practical equivalence. Age erscheint 2 x, wollen wir das so ? Sind 2 Farben beabsichtigt (grau und blau) und wir müssten noch sagen, wofür der graue Balken steht)

We state this plainly because it contradicts our initial expectations. The predictor that education can modify is the weakest in the model, and the three strong predictors are all forms of experiential exposure acquired outside the curriculum. The dose–response analysis points in the same direction: across the four course levels, self-assessed competence rises from 3.33 to 4.92 on the six-point scale, whereas formal willingness rises only from 71.0% to 83.2% (see Multimedia Appendix 8).

### Public Health Roles and Closing Items

When asked about possible contributions to public health services, respondents most often endorsed vaccination campaigns (67.7%) and the compilation of simple health situation reports (50.6%), ahead of citizen information (44.5%), information hotlines (33.5%), infection control assistance (33.0%), and surveillance support (32.5%). When asked about the single most important measure to improve integration, 29.9% named binding integration into the curriculum, 27.3% named attractive voluntary additional offerings, and 16.7% named financial and legal incentives. Participation in the survey changed interest in the topic somewhat for 54.9% of respondents and markedly for 12.0%, who stated that they would actively seek further information on the topic.

## Discussion

### Principal Results

Nearly two decades after curricular recommendations were issued, the largest interprofessional survey of its kind indicates that disaster medicine remains largely absent from health professions education in Germany. A third of respondents had encountered nothing at all, and where content had been delivered, it was mostly not retained: across twelve topics, the proportion able to retrieve content confidently never exceeded 23.9%, and for three operationally central topics, a majority had met the material and lost it. More than half had never participated in any practical major incident exercise, and where exercise experience existed, it came predominantly from civil protection organizations rather than educational institutions.

In our view, three further findings are more consequential than the deficit itself because they concern how educational interventions should be designed rather than whether they are needed.

### Delivery Is the Wrong Indicator

Curriculum reviews conventionally ask whether a topic is covered. Our data suggest that this indicator is close to being uninformative in this content area. The delivery–retention gap reached 47.3 pp, and it was largest precisely for the topics that would be immediately action-guiding under deployment conditions: ethical prioritization, decontamination and self-protection, and structures of civil protection. A faculty member reporting that these topics are covered would be reporting accurately and would nonetheless be describing a curriculum whose graduates cannot use the content of the course.

Two mechanisms are plausible, but we cannot distinguish between them here. The material may be delivered in a form and at a spacing that does not support retention, typically a single lecture embedded in an unrelated module without subsequent retrieval practice or assessment. Alternatively, it may be retained in principle but not experienced as retrievable because respondents have never had to apply it and therefore lack the confidence that grounds a claim of retrievability. Both mechanisms point in the same educational direction, toward distributed practice, retrieval, and application, but they differ in whether the deficit is one of knowledge or self-efficacy. Distinguishing them requires an objective knowledge assessment, which our design does not provide and which we regard as the single most useful next step.

### Competence in This Field Has More Than One Dimension

The competence battery showed high internal consistency yet retained three factors, and the loading pattern was unambiguous: 18 acute-technical items formed a strong first factor, while four relational and nursing items were almost outside it. Differential item functioning analysis then shows a complete directional reversal by prior civil protection engagement, with higher acute-technical confidence and lower relational confidence without a single ambiguous item. We read this as a substantive, rather than psychometric, result. The socialization that civil protection engagement provides is powerfully effective for technical emergency skills and does not transfer to, and may compete with, relational and nursing competencies. This has direct design implications: in an interprofessional preparation program, relational and nursing competence must be treated as distinct learning objectives with their own outcomes and assessments, not as something that accrues alongside clinical training. This has methodological implications for the field. A monoprofessional study of medical students would not have detected this pattern because instruments developed within medicine rarely include personal care, assistance with eating, or providing orientation as competence. Multidimensionality is visible only when the instrument spans professions.

### Curricular Exposure and Experiential Exposure Are Different Things

Our initial expectation, shared with most calls for curricular expansion, was that curricular exposure would predict willingness. In the adjusted model, it did so only weakly, with an odds ratio of 1.06 per standard deviation and an interval inside the prespecified region of practical equivalence, while active civil protection engagement carried an odds ratio of 2.49 and practical exercise experience 1.78. The dose–response pattern was consistent: course level substantially raised self-assessed competence and only modestly raised willingness.

The most plausible alternative explanation is self-selection, and the cross-sectional design cannot exclude it: people already disposed to serve join response organizations and attend exercises. We regard this as at least as likely as a formative interpretation and state so. What the data establish, whichever direction the association runs, is that curricular exposure and experiential exposure are not interchangeable, and that the component education controls are the weaker of the two.

This does not argue against curricular reform. It argues against justifying curricular reform by its expected effect on willingness. The case for teaching disaster medicine rests on competence and safety, and our competence data strongly support this case. Willingness in this population is already high and appears to be limited by structural rather than motivational factors.

### The Barriers Are Structural

Motivation in this population is intrinsic; practical competence and social contribution outrank financial and academic incentives. These obstacles are structural, including insurance, time, and legal ambiguity. Academic credit ranked last among the eight framework conditions, below insurance and certification, and supervision by experienced instructors ranked first at 95.6%. However, credit and certification are precisely the levers that curriculum designers control most directly, and supervision is the scarcest and most expensive resource available.

We believe that this tension cannot be resolved by design alone. The condition that the target group considers most important is also the one that the deploying organizations, in the companion analysis of the same project, name as their second strongest concern in the form of supervision burden ^13^. An offer that promises experienced supervision without securing the personnel to provide it will disappoint the dimension that matters most to its participants.

The implications extend beyond education. To design training that is operationally meaningful, the wider complex adaptive system in which future responders will act must be considered. From a systems thinking perspective, no single actor controls the whole system in a complex crisis; deployable capacity therefore emerges from the interaction of willing individuals, training structures, supervision, legal frameworks, and deploying organizations rather than from individual willingness alone. Such capacity depends on relationships, shared language, trust, transparent decision-making, and practiced adaptation mechanisms that enable multiple actors to maintain fairness and continuity under pressure. System performance should therefore not be judged by how well any single organization performs in isolation, but by whether the interacting parts of the system collectively maintain essential functions, adapt to disruption, and sustain effective action over time ^21^. Experience from the regional COVID-19 response showed how this translates into practice: rapid mobilization depended on robust mandates, established networks, and structures for onboarding and securing the commitment of additional actors ^22^. Subsequent lessons learned from pandemic disaster management extended this principle to volunteers and spontaneous helpers, who can constitute an important workforce resource but require pre-established structures for integration and onboarding ^23^.

Finally, the time budget was compatible with meaningful provision: 75.9% could accommodate at least one weekend per half-year or the equivalent, and only 1.4% ruled out any additional time spent. The constraint is not availability but rather the format and conditions.

### Implications for Curriculum Design

#### 1. Prioritize by want–deficit, not by coverage

The three topics in which demand and deficit coincide most strongly are trauma care under resource scarcity, triage, and the structures of civil protection. These have the best joint justification for inclusion in this study. Self-protection should be added on external grounds: it carries the second largest delivery–retention gap, and in the companion analysis, the deploying organizations ranked it first among desired training contents while showing the largest gap between their requirement and students’ self-assessed confidence ^13^.

#### 2. Design for retention, not delivery

Given the size of the delivery–retention gap, single-exposure formats are unlikely to be sufficient regardless of their content. Distributed practice with retrieval opportunities and applications, which the format preferences of this population already favor, should be the default assumption of any implementation. Evidence from a five-year evaluation of a curricular elective module in disaster and deployment medicine supports the feasibility of sustained mixed-format delivery at the university level ^24^.

#### 3. The multidimensional structure and reversal by prior engagement mean that relational and nursing

competence will not accrue as a byproduct of clinical or civil protection training. An interprofessional program requires its own outcomes and assessments.

#### 4. The format must match the stated budget and conditions

One weekend per half-year in a hybrid design with a high practical component and joint exercises with response organizations matches the preference and budget of this population. Making the offer free of charge and providing experienced supervision mattered more to participants than credit or certification.

#### 5. Do not justify the program by its effect on willingness

Willingness is already high, largely decoupled from competence, and limited by insurance, time, and legal ambiguity. These are not curricular problems, and a program sold on its motivational effect will be measured against an outcome that it is unlikely to move.

### Comparison With Prior Work

Our findings reproduce and extend this established picture. Wunderlich et al. surveyed 992 German medical students nine years after the approval of the concept curriculum, and Kasselmann et al. surveyed 25 of 36 medical faculties, both of which concluded that disaster medicine remains underrepresented and institutionally heterogeneous ^7,8^. National surveys from the Netherlands and Italy reported similar findings for medical students ^9,10^. A systematic review of student volunteering found that 36.8% of 76 medical school curricula included disaster teaching, and that where it was included, students received only two to six hours ^12^. The proportion of participants reporting no deliveries was of the same order. For nursing, a recent systematic review of 19 studies concluded that simulation-based and scenario-driven approaches produce the most consistent gains in self-efficacy, which is consistent with the format preferences observed in this study ^11^.

The extension lies in three directions: The interprofessional breadth shows that the deficit is not a peculiarity of medical curricula and that its size differs substantially between professions, with the therapeutic and psychosocial professions lagging behind in terms of exposure and competence. The delivery–retention distinction shows that the problem is larger than the delivery figures suggest. The adjusted model provides, as far as we can establish, the first test of the assumption that curricular exposure drives willingness, with a result that does not support this assumption.

The Italian blended learning program, which reached approximately 2,500 students across 37 faculties over six years through student-led courses and e-learning, remains the most instructive implementation model in the literature ^25^. Its format profile corresponds closely to the preferences expressed by our respondents, which strengthens the case for it as a template, rather than a national exception. Qualitative work on medical students in hospital emergency preparedness after the COVID-19 pandemic points in the same direction and adds the perspective of receiving institutions ^26^.

### Limitations

The principal strengths are the size and interprofessional breadth of the sample, which permit group comparisons and adjusted modeling simultaneously, the separation of delivery from retention, and the prespecification of the predictor set, the region of practical equivalence, and the analysis code, which was frozen before application to the actual data.

- Delivery and retention were self-reported by the participants. Self-assessed retrievability is not a knowledge test and may be biased in either direction, which cannot be determined from these data. Therefore, all statements are about self-reported delivery and retention.
- The cross-sectional design did not permit causal statements. The association between exposure and willingness is equally compatible with formation and self-selection, and we cannot separate them from each other.
- The sample was self-selected without a sampling frame; therefore, no response rate could be calculated in the strict sense. The proportion of prior civil protection engagement was a selectivity indicator and was examined in the prespecified subgroup analysis. The weighting of the official marginal distributions foreseen in the analysis plan could not be executed because the required external reference distributions were unavailable.
- Duplicate participation was prevented by browser-based cookies; however, because IP addresses were not stored and no access tokens were used, repeat entries from a different browser or device could not be excluded; this is an accepted trade-off of the anonymous design.
- Willingness showed a ceiling effect that compressed the variance and may have attenuated group differences. The ordinal model uses the full response distribution partly for this reason; however, the constraint remains unchanged.
- The instrument was not externally validated, and the psychometric properties reported here are the first determination, labeled exploratory.
- All results concern stated willingness, not observed behavior; the intention–behavior gap is well documented and has been observed in this specific population ^12,27,28^.
- The occupational groups differ in size, and the three smallest groups comprise fewer than 400 respondents each; therefore, estimates for these groups are less precise than the overall figures.
- This is a single-country study within one educational and civil-protection system. The questions are likely to be generalizable, although the answers may not be.

### Directions for Further Research

Three next steps appear particularly important. First, aAn objective knowledge assessment alongside self-report would establish whether the retention deficit is due to knowledge or self-efficacy, which determines the educational response. Second, a longitudinal design following students through curricular intervention would separate the formative interpretation of the exposure–willingness association from the selection interpretation. Third, a study relating the multidimensional competence structure to observed performance in a mixed interprofessional exercise would test whether the dimensions identified psychometrically correspond to distinguishable performance in practice.

### Conclusions

Almost two decades after disaster medicine was recommended for inclusion in German health profession curricula, a third of students and trainees have encountered none of it, and most of what has been delivered cannot be retrieved from memory. The deficit spans all seven occupational groups examined and is largest in the therapeutic and psychosocial professions, which are not covered in the literature.

Nonetheless, the willingness to serve is high and largely decoupled from competence. Qualification and recruitment are therefore separate problems requiring separate instruments: education should be justified by competence and safety, whereas recruitment depends on insurance, time, and legal clarity, none of which are curricular issues. Where education can act, it should prioritize the coincidence of demand and deficit rather than coverage, design for retention rather than delivery, and treat relational and nursing competence as distinct learning objectives rather than as byproducts of clinical or emergency training.

## Declarations

### Data Availability

The anonymized data set, the codebook, both survey instruments, and the complete analysis code generated and analyzed during this study will be deposited in a public repository upon publication (data under CC BY 4.0, code under the MIT license). Until then, the data are available from the corresponding author on reasonable request. Free-text fields are screened for potentially identifying information before release, and cell counts below 5 are aggregated.

### Authors’ Contributions

KFF, MRi, and TZ contributed equally and share first authorship. PR, ACF, and MvdF contributed equally and shared the last authorship.

Conceptualization: KFF, MRi, TZ, VE, RW, PB, PF, ACF and MvdF. Methodology: KFF, MRi, TZ, BE, RW, PB, AFo, PR, and MvdF. Software: TZ, PR. Formal analysis: KFF, TZ, PR. Validation: MRi, BE, RW, AFo, KM, MRe, and PR. Investigation: KFF, BG, FF, VE, AK, ES, LOM, WZ, PF, KB, OF, and ACF. Resources: FF, PF, KB, DD, and OF. Data curation: KFF, TZ, BG, AK, and ES. Writing – original draft: KFF, TZ, MRi, PR, and MvdF. Writing – review and editing: All authors. Visualization: TZ and KFF. Supervision: KFF, MRi, DD, PR, ACF, and MvdF. Project administration: KFF, PR, ACF, and WZ. Funding acquisition: KFF and DD. All authors have read and approved the final manuscript.

## Acknowledgments

We thank the expert panel of the instrument consenting process, the pretest participants, and the institutions that forwarded the survey invitation, subject to their consent to be named in this manuscript.

## Companion Papers

Three further analyses of the same data from the KOMPaKT112-MED/ORG study are presented here. The task-level alignment between students and trainees and the organizations that would deploy them was reported separately [13]. An analysis of organizational respondents’ perceived deployment needs, competence requirements, and governance expectations, and a mixed-methods analysis of the free-text justifications are in preparation. No endpoints were reported as primary results in any of these studies.

